# Modelling the potential influence of human migration and strain mutation on Ebola virus disease dynamics

**DOI:** 10.1101/2022.05.29.22275751

**Authors:** Sylvie Diane Djiomba Njankou, Farai Nyabadza

## Abstract

Migration of infected animals and humans, and mutation are considered as the source of the introduction of new pathogens and strains into a country. In this paper, we formulate a mathematical model of Ebola virus disease dynamics, that describes the introduction of a new strain of ebolavirus, through either mutation or immigration (which can be continuous or impulsive) of infectives. The mathematical analysis of the model shows that when the immigration of infectives is continuous, the new strain invades a country if its invasion reproduction number is greater than one. When the immigration is impulsive, a newly introduced strain is controllable when its reproduction number is less than the ratio of mortality to the population inflow and only locally stable equilibria exist. This ratio is one if the population size is constant. In case of mutation of the resident strain of ebolavirus, the coexistence of the resident and mutated strains is possible if their respective reproduction numbers are greater than one. Results indicate that the competition for the susceptible population is the immediate consequence of the coexistence of two different strains of ebolavirus in a country and this competition is favourable to the most infectious strain. Results also indicate that impulsive immigration of infectives when compared to continuous immigration of infectives gives time for the implementation of control measures. Our model results suggest controlled movements of people between countries that have had Ebola outbreaks despite the fact that closing boundaries is impossible.

## 1 Introduction

More than 20 Ebola Virus Disease (EVD) outbreaks hit the African continent in recent decades and different ebolavirus strains have caused these outbreaks. There are five different strains of ebolaviruses and the most devastating are the Zaire ebolavirus strain, the Sudan ebolavirus strain and the Bundibugyo ebolavirus strain [1]. Among all the theories explaining how a Central African virus moved to West Africa, one of them supports the idea that migrating bats that can fly over hundreds of kilometres might have brought the virus there [3]. Migration of the natural reservoir of ebolavirus in this case is a key element in the transportation of the Zaire ebolavirus strain within different parts of the continent. Economic activities such as hunting or mining have increased the contacts between humans and infected animals such as bats, chimpanzees or monkeys living in tropical forests [4, 5]. An eventual mutation of the Zaire ebolavirus strain was suspected as well because of the large death toll of the outbreak. The African tropical forest that hosts Ebola viruses natural reservoirs ranges from East to West Africa [6]. The Zaire, Sudan and Bundibugyo ebolavirus strains have been linked to the animal population during the past outbreaks and this might explain the fact that different species have hit several countries already [4, 5]. The Democratic Republic of Congo (DRC) generally hit by the Zaire strain was affected by the Bundibugyo strain in 2012, showing the possibility of one region being affected by a different strain. Uganda naturally hosts the Bundibugyo ebolavirus strain and was hit by the Sudan strain in 2000 and 2011 − 2013. Besides, DRC, Uganda and Sudan share common boundaries [1].

Movements within the African continent are motivated by labour and livelihoods, social and familial connections, cultural ceremonies, disasters and conflicts [7]. Eastern and Western Africa have a long history of labour migration between and within countries to plantations (cotton and coffee in Uganda, cocoa and coffee in Ivory Coast) or to mines (in DRC and Uganda) during the appropriate seasons [8]. Pastoralist communities in Kenya, Tanzania, and Uganda for example, move to feed their animals [8]. In West Africa, the labour force from Mali, Burkina Faso and Guinea frequently move to Ivory Coast to harvest cocoa [9, 10]. This type of migration is often seasonal and motivated by the climate change or the harvesting period and is somehow similar to impulsive migration. Tourists’ movements are also a typical example of impulsive migration and in Uganda for example, intra-African migrant birds and the tourists coming to see them arrive in July and start leaving in December [11]. Borders and boundaries in West Africa are highly porous and make it near impossible to track people’s movements [7]. Fear, stigmatization or the non-access to close Ebola health care centers unexpectedly increased populations’ mobility during the outbreak of 2013 − 2016 [7]. Given that different strains exist in the West and Central African regions, with free movement of persons, the potential of new strain exportation exists. This phenomenon can be viewed as migration of infectives in mathematical modelling of communicable diseases and we consider the potential of such migration in the spread of EVD.

Mathematical models of EVD that include migration of individuals have been formulated and analysed by several authors, Valdez *et al* [13], a stochastic model for Ebola, Kramer *et al* [14], spatial spread of EVD, Brauer and Van Den Driessche [15], immigration of infectives in an HIV/AIDS model, in [15], HIV/AIDS transmission in a prison and Tripathi et al [16], HIV/AIDS with immigration of infectives, treatment and time delay.

This paper considers the possibility of strain importation through human migration and strain mutation, and therefore present a possibility that is highly likely in the distant future. We thus formulate a model of EVD with a resident strain that either mutates or is invaded by a new strain of ebolavirus and evaluate the potential influence of strain importation or mutation on the disease dynamics. If the new strain is a mutant of the resident strain, then we have a scenario where the possibility of an invasion by the new strain is possible. If the new strain is imported from a different region, then we consider the cases where we have a continuous or an impulsive migration of infectives. We assume that movements of populations on the African continent could lead to the importation new strains of EVD into a country already affected by different strains.

This work is arranged as follows: he model formulation is given in Section 2, followed by the analysis of the models with 1) continuous migration of infectives in Section 3, 2) mpulsive migration of infectives in Section 4 and 3) with mutation of the resident strain in Section 5. Numerical simulations are done in Section 6 and concluding remarks in Section 7.

## 2 Model formulation

We formulate a model of EVD with two strains, strain 1 (EVD1) and strain 2 (EVD2). Strain 1 is considered to be the resident strain while strain 2 is either an imported or a mutant strain. In a given region or country, we assume that susceptible individuals, at any time *t*, denoted by *S*(*t*), are recruited at a constant rate Λ either through births or immigration. Susceptible individuals can be infected by either strain 1 or strain 2 at a time. We thus do not assume co-infection by different strains. Susceptible individuals infected by strain *i* (*i* = 1, 2), move to compartment *I*_*i*_ and can transmit EVD. An infectious individual can either die of the disease or recover. Those that die of strain *i* are denoted *D*_*i*_ and those that recover are assumed to belong to the class *R* that is independent of the strain they were suffering from. Susceptible, infected and recovered individuals die naturally at a rate *µ* with the infected dying at rates *φ*_1_ and *φ*_2_ from strain 1 and 2 respectively. Individuals are assumed to recover from strain 1 and strain 2 at a rates *α*_1_ and *α*_2_ respectively. We also assume that recovered individuals acquire immunity during the modelling period. The deceased are assumed to contribute to infection and the force of infection for the respective strains is *λ*_*i*_ = *β*_*i*_ (*I*_*i*_ + *η*_*i*_*D*_*i*_) for *i* = 1, 2 where *η*_*i*_ is a parameter that measures the relative infectivity of the deceased compared to the infected. To model migration that leads to the importation of strains, we consider a constant migration rate *σ* in which a proportion *p* is infectious and the remainder is susceptible. Ebola epidemics are usually short and we do not assume migration of the recovered and deceased during an epidemic. It is important to note that the constant Λ incorporates migrants and births in a community. The corpses of the deceased are disposed at rates *ρ*_*i*_ depending on the strain that caused the death. The flow of the individuals between compartments is shown in Figure 1.

**Figure 1:**
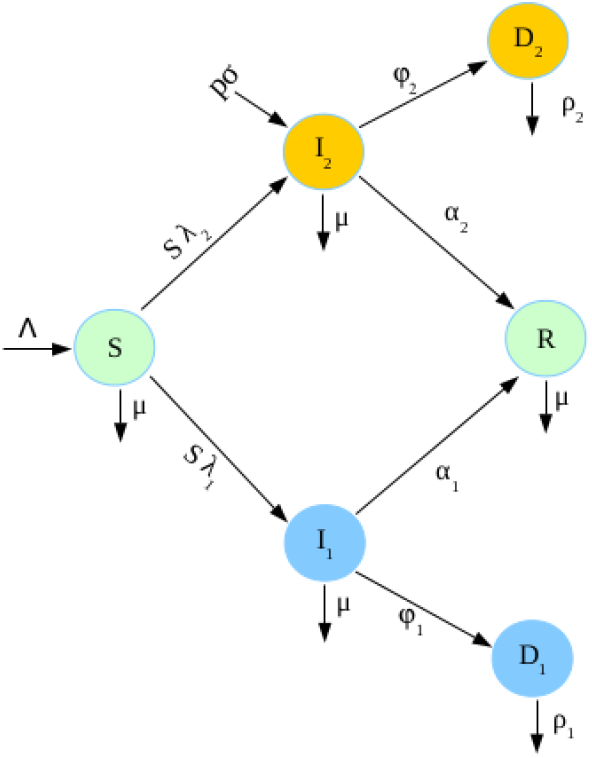
Flow chart diagram of the model with migration of infectives

The differential equations that model the described disease dynamics are

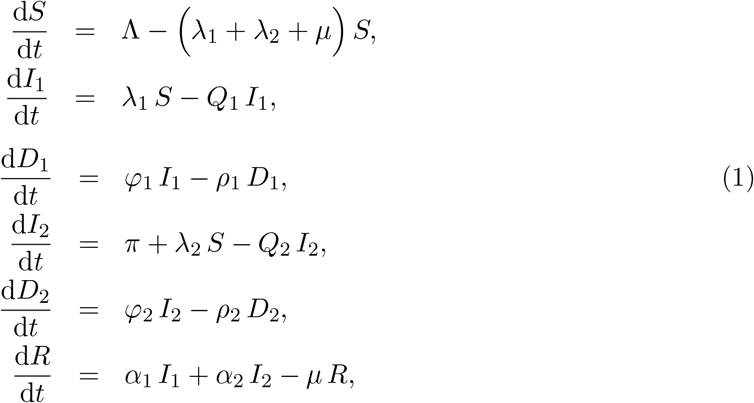

with initial conditions

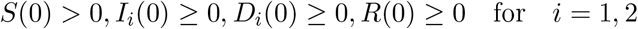

and where *Q*_1_ = *µ* + *α*_1_ + *φ*_1_, *Q*_2_ = *µ* + *α*_2_ + *φ*_2_. We define Λ = *θ* + (1 − *p*) *σ* and *π* = *pσ* where *θ* is the recruitment rate of susceptibles through other means, other than immigration. We consider the equation for *R* to be redundant. To analyse (1), we consider three cases: first, a case with a constant immigration of infectives who come with strain 2, second a case in which we have impulsive migration, and third a case in which we have a mutant strain 2 that comes from strain 1 with the recovered class considered as redundant.

## 3 Model of Ebola dynamics with continuous immigration of infectives

We consider the case where individuals infected by strain 2 of ebolavirus are constantly recruited into a country already affected by strain 1 of ebolavirus. The flow between the compartments of the model representing Ebola dynamics in this case is given by

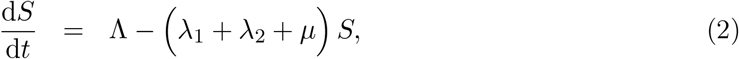

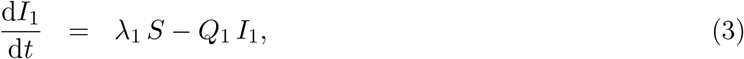

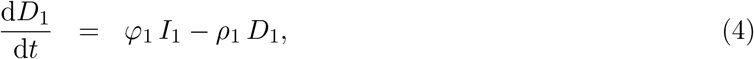

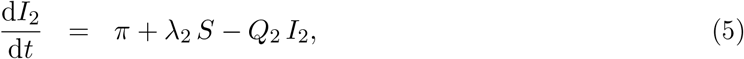

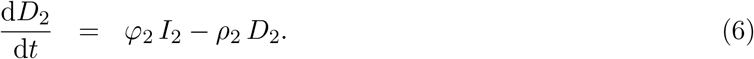

### 3.0.1 Existence and positivity of solutions

System (2)-(6) makes biological sense if its solutions exist and are positive in an invariant region.

#### Theorem 3.1.

The invariant region is given by

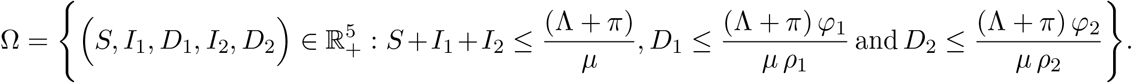

*Proof*. Let us set *M* (*t*) = *S*(*t*) + *I*_1_(*t*) + *I*_2_(*t*). Adding equations (2), (3) and (5) yields

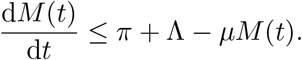

Solving the above differential equation and using the Gronwall inequality yield 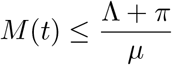 Similarly, since 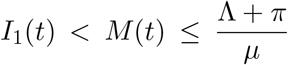, equation (4) yields 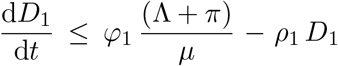 and Gronwall inequality gives 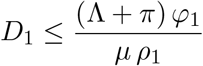.

Since 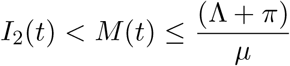, equation (6) yields 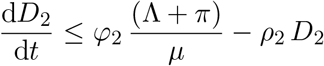 and similarly we obtain 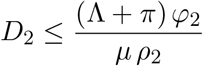.

#### Theorem 3.2.

*All the solutions of the system* (2)*-*(6) *are non-negative for non-negative initial conditions*.

*Proof*. We set *A*(*t*) = *λ*_1_(*t*) + *λ*_2_(*t*) + *µ*. Solving equation (2) for *S*(*t*) yields.

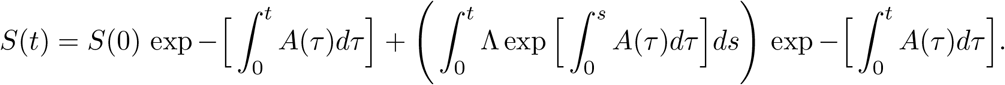

So, *S*(*t*) ≥0 for all *t* ≥0 whenever *S*(0) ≥0. System of equations (3)-(6) can be rewritten as 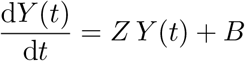 where

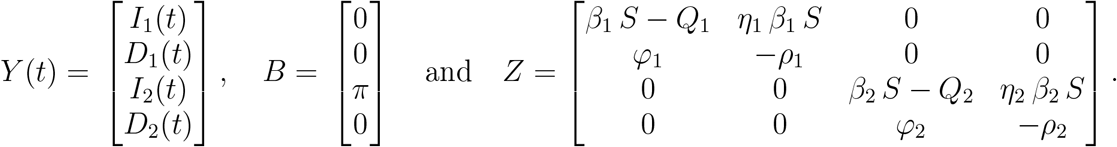

Since all the off diagonal elements of *Z* are non-negative, *Z* is a Metzler matrix and *Y* is monotone and positive, see [19, 20]. So, 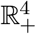 is invariant under 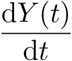 and *Y* (*t*) is non-negative.

### 3.0.2 Reproduction number

The reproduction number *R*_0_ is calculated by using the next generation matrix method, see [21]. We find *R*_0_ = max {*R*_1_, *R*_2_} where

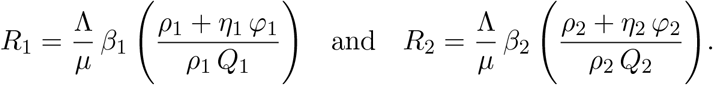

The condition *π* = 0 is a necessary condition in order to reach the total absence of EVD. In this case, global stability of the equilibrium point is not guaranteed as long as infected immigrants continue to move into the country. This emphasizes the complexity of the control of EVD in the African setting where road boundaries particularly, are most of the time porous and migration is not always well controlled.

### 3.0.3 Strain 2 free equilibrium

In the absence of EVD2, the reproduction number is *R*_1_ and the endemic equilibrium is 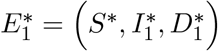 where

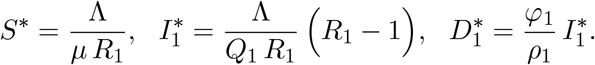

#### Theorem 3.3.

The endemic equilibrium 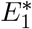 exists for *R*_1_ *>* 1. Before the invasion of strain 1 by strain 2, 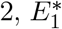 is globally asymptotically stable. When the invasion occurs, 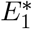 is locally stable.

*Proof*. Strain 2 can invade strain 1 when the latter is at equilibrium and the invasion reproduction number of strain 2 denoted by 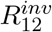 is computed using the next generation matrix method for 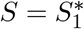 and *I*_2_ = *D*_2_ = *π* = 0. We obtain

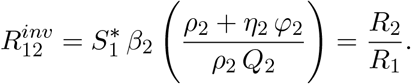

The use of the next generation matrix method to compute 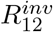 guarantees the local stability of the equilibrium point 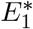. The proof of the global asymptotic stability of 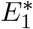 before the invasion is as follows: we set *F*_1_ as the Lyapunov function with

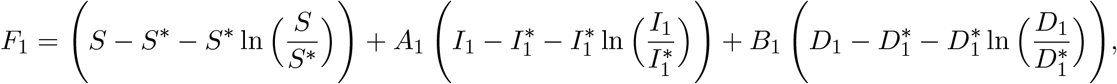

where *A*_1_ and *B*_1_ are positive constants to be calculated with 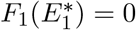. The right hand side of system (2)-(6) at equilibrium yields

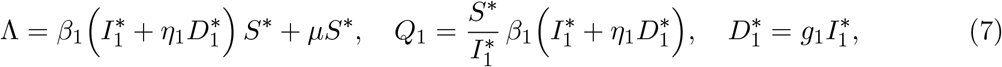

Where 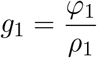. 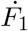 is the derivative of *F*_1_ with respect to time and is given by

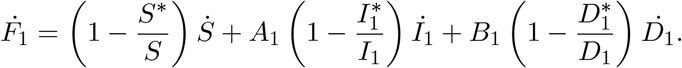

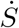 is obtained from equation (2), 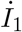 is obtained from equation (3) and 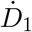 is obtained from equation (4). We then have

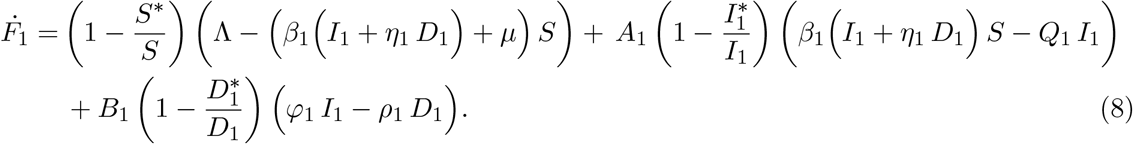

We set

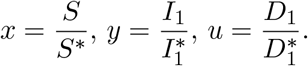

Using the expressions in (7) and *x, y, u* into (8) yield

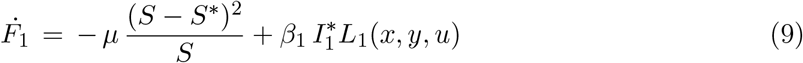

where

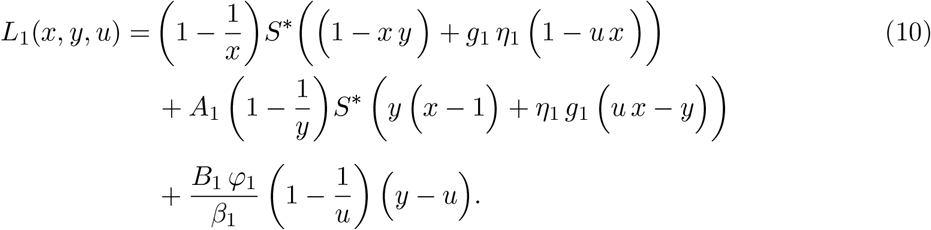

Equation (9) implies 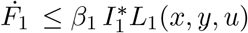 since 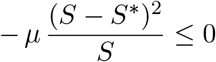.

Expanding the expression of *L*_1_(*x, y, u*) from system (10) and grouping the coefficients with the same variable and sei the terms with non-negative coefficients to zero gives

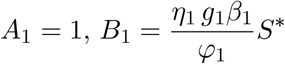

and

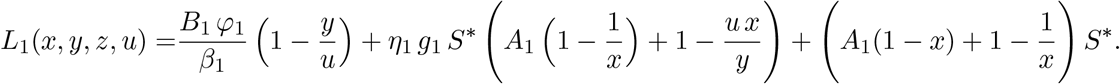

So, for *x* = *y* = *u* = 1, *L*_1_ is negative and equal to zero. So, *L*_1_ ≤ 0 for (*S, I*_1_, *D*_1_) ∈ Γ where

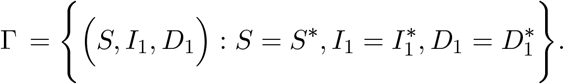

By LaSalle’s invariance principle, see [27], 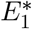 is globally asymptotically stable on Ω.

In the country, where EVD1 already exists, immigration of individuals infected by EVD2 may lead to an invasion and the invasion reproduction number is *R*_12_. If *R*_1_ *> R*_2_, then EVD1 is spreading faster than EVD2 which may vanish at some point. *R*_1_ *<* 1 is necessary to stop EVD1 but not EVD2. *π* = 0 is necessary for the total eradication of EVD2. If *R*_2_ *> R*_1_, then EVD2 totally invades the country and reducing *R*_2_ to values less than one will help to limit the spread of strain 1 since *R*_2_ *<* 1 implies *R*_1_ *<* 1 in this case. So, in order to stop the invasion, immigration of individuals infected by EVD2 should be prohibited and other control measures such as quarantine and hospitalisation should be introduced to limit the spread of EVD1 and EVD2 in the population. When EVD1 is the only strain of EVD existing in the country, its eradication is easier and health authorities should focus on limiting its spread among the population and encourage mostly movements within the country.

### 3.0.4 Coexistence equilibrium

The coexistence of the two strains leads to the endemic equilibrium 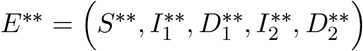 where

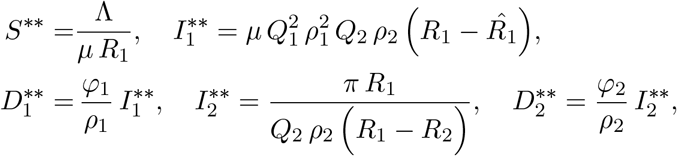

with 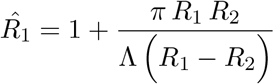

So EVD1 and EVD2 can coexist when EVD1 is dominant. *R*_1_ *>* 1 is not enough for EVD1 to coexist with EVD2. More individuals must be infected by EVD1 for both strains to coexist. The coexistence of the two strains is conditioned by 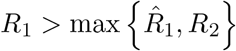. Figure 2 shows the region of existence of *E*^∗∗^ when *R*_1_ and *R*_2_ are varied.

**Figure 2:**
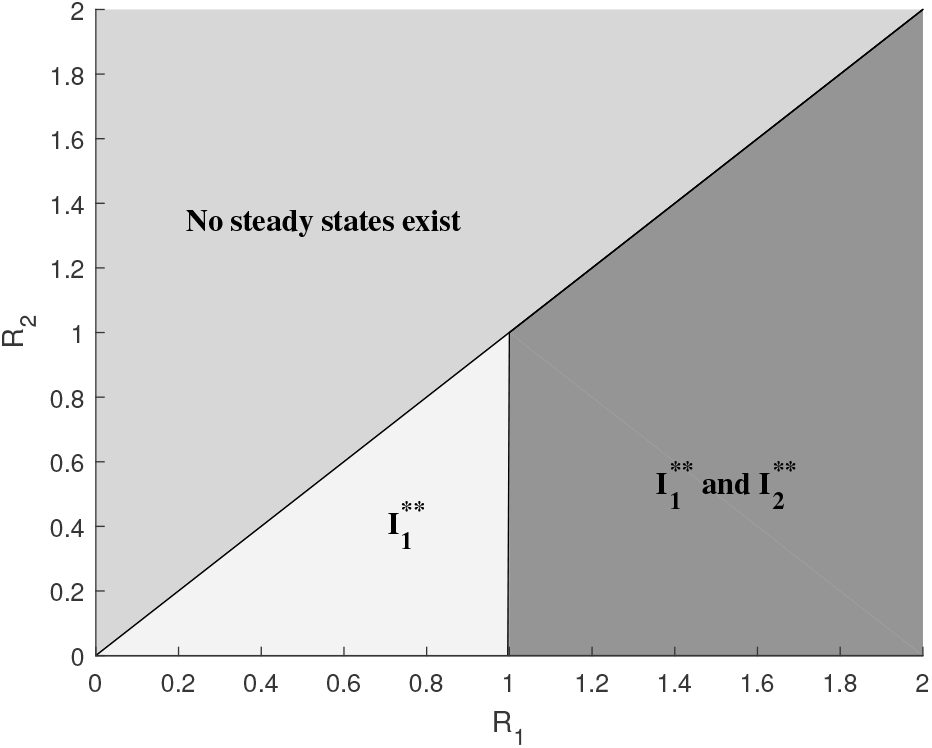
Region of existence of 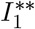 and 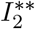

We observe in Figure 2 that there is no region of existence of 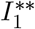 only and 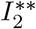 only exists when *R*_2_ *<* 1. This is due to movement of infectives which always guarantees the presence of individuals infected by EVD2 and suppresses the case where only individuals infected by EVD1 exist. The constant immigration of infectives in this case makes EVD control more difficult since reducing *R*_2_ to values less than one is not enough to reach the DFE.

#### Theorem 3.4.

The endemic equilibrium *E*^∗∗^ is locally asymptotically stable.

*Proof*. To prove the local stability of *E*^∗∗^, we set *R*_0_ = *R*_1_ since *R*_1_ *> R*_2_ is a necessary condition for the existence of *E*^∗∗^ and *R*_0_ = max {*R*_1_, *R*_2_}. In order to describe the local stability of the endemic equilibrium, we will use the theorem, remark and corollary which are based on the Centre Manifold Theory [22].

We set *ϕ* = *β*_1_ as our bifurcation parameter, so that for

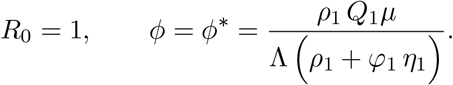

Following [22], the Jacobian matrix 𝒥 of the linearised system (2)-(6) at the DFE *E*^0^ and for *ϕ = ϕ\** is given by

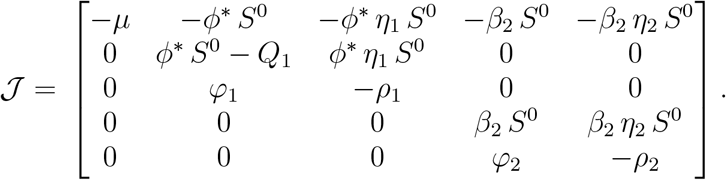

Zero is a simple eigenvalue of 𝒥. The left eigenvector of 𝒥, *V* = (*v*_1_, *v*_2_, *v*_3_, *v*_4_, *v*_5_) and the right eigenvector *W* = (*w*_1_, *w*_2_, *w*_3_, *w*_4_, *w*_5_) ^′^, after some algebraic manipulations are given by

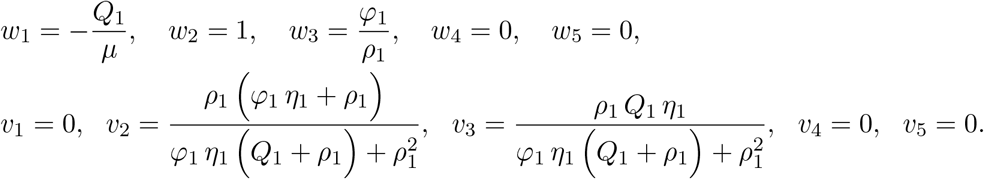

Besides, we notice that for *j* = 2, 3, 4, 5, *E*^0^(*j*) = 0 and *W* (*j*) is non-negative. So remark 4 in [22] is verified. Using the formulas defined in [22], we compute the constants *a* and *b* and find

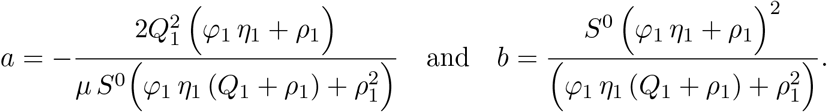

The direction of the bifurcation is determined by the signs of *a* and *b*. Obviously *b >* 0 and *a <* 0 indicating that *E*^∗∗^ is locally asymptotically stable and the bifurcation is forward.

## 4 Model of Ebola dynamics with impulsive immigration of infectives

Impulsive differential equations (IDE) have been produced since 1990 and describe the dynamics of evolving processes subjected to short-term perturbations that act instantaneously or in the form of impulses [23]. We consider in this case that EVD2 is introduced into a population already affected by EVD1 in the form of impulses at specific times *t*_*k*_, *k* = 1, 2, …, *m* with *m >* 0. During the specific times *t*_*k*_, the boundaries of the country affected by strain 1 are opened and groups of individuals move in. Individuals infected with EVD2 are recruited at a rate *π*. The system of IDE describing the flow of individuals is given by

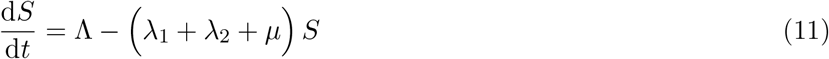

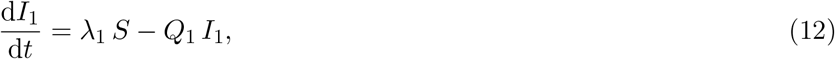

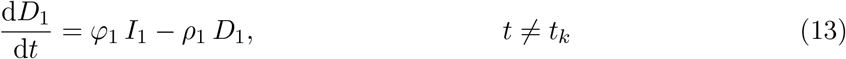

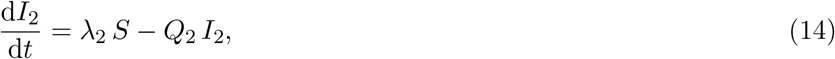

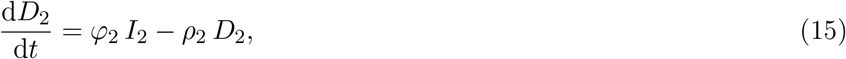

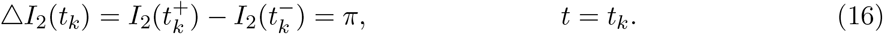

Considering only the state variables affected by the impulse, we obtain system (17)-(18).

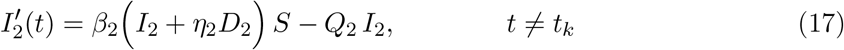

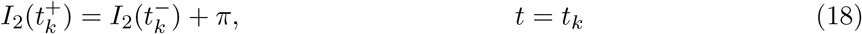

where *t*_*k*+1_ *> t*_*k*_. We assume in this case that there is no individual infected by EVD2 before the first impulse, so 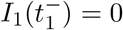 and we set 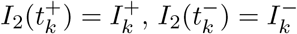.

### 4.0.1 Positivity of solutions

For *t* ∈ (*t*_*k*_, *t*_*k*+1_], system (11)-(16) is equivalent to (2)-(6) where *π* = 0 and whose solutions has already been proven non-negative in Theorem 3.1. Between two consecutive impulses, solutions of system (11)-(16) are positive and Ω remains the invariant set.

### 4.0.2 Existence and uniqueness of solutions

#### Theorem 4.1.

Solutions of system (11)-(16) exist in the sets (*t*_*k*_, *t*_*k*+1_] × Ω and are unique for each initial condition (*t*_0_, *x*_0_) ∈ R_+_ × Ω. Besides, each solution *φ* : (*α, β*) → R^*n*^, *α, β* ∈ Z_+_, *α < β, β*≠ *t*_*k*_, is continuable to the right of *β*. The general expression of the maximal solution of (11)-(16) is given by 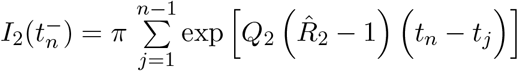 where 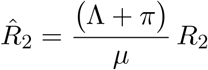.

*Proof*. The proof is based on the Theorem in [24] stipulating the conditions for the existence and uniqueness of the solutions of a system of non linear IDE with fixed moments of impulses. The Theorem states that given an IDE,

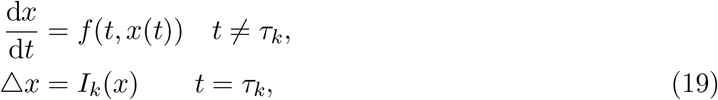

where *τ*_*k*_ *< τ*_*k*+1_ (*k* ∈ Z) and 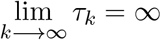.

Let the function *f* : R × Ω −→ R^*n*^ be continuous in the sets (*τ*_*k*_ × *τ*_*k*+1_] × Ω. For each *k* ∈ Z and *x* ∈ Ω, suppose there exists the finite limit of *f* (*t, y*) as (*t, y*) −→ (*τ*_*k*_, *x*), *t > τ*_*k*_.

Then, for each (*t*_0_, *x*_0_) ∈R Ω, there exists *β > t*_0_ and a solution *φ* : (*t*_0_, *β*) R^*n*^ of the initial value problem (19). Moreover, if the function *f* is locally Lipschitz continuous with respect to *x* in R × Ω, then this solution is unique. Besides, if 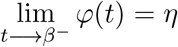 and *η* ∈ Ω when *β*≠ *τ*_*k*_, then solution *φ*(*t*) is continuable to the right of *β*. The general solution is in the form

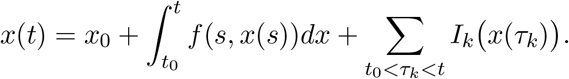

The right hand side of system (11)-(15) is bounded and locally Lipschitz in the sets (*τ*_*k*_ ×*τ*_*k*+1_]×Ω, for *k >* 0 and *α, β* ∈ (*τ*_*k*_ × *τ*_*k*+1_] with *β*≠ *τ*_*k*_. 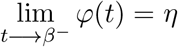 belongs to Ω since Ω is positively invariant and we can conclude that the solutions of system (11)-(16) exist and are continuable to the right of *β* ≠ *τ*_*k*_. The non linearity of the system of equations (11)-(15) makes it difficult to find its algebraic solution. Instead, we give the expression of the maximal solution that does not contain this non linearity.

### Evaluation of the maximal solution

From the invariant set 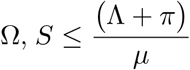 and 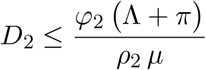. Besides,*D*_*2*_ *< D*_2_ *I*_2_ and equation (17) is maximised. We obtain for *t*≠ *t*_*k*_

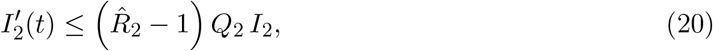

where 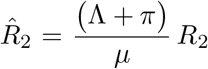. We solve the equality corresponding to equation (20) during a single impulsive cycle, 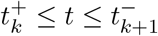 and obtain

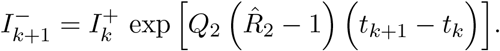

From equation (18), 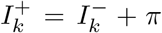 and this implies that 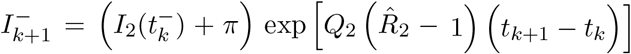. Since 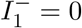 we can write

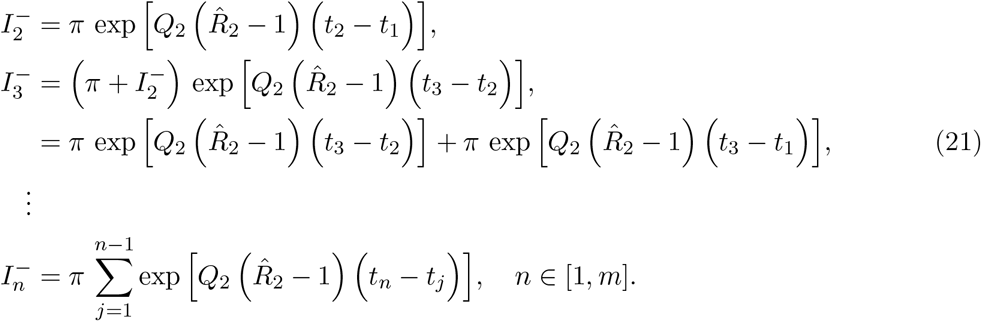

Adding all the equations of the system (21) yields

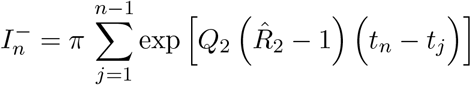

and 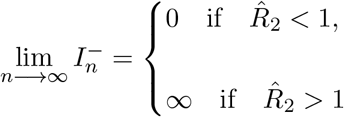 since 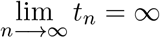.

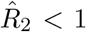 implies 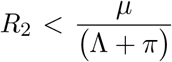. So, reducing the reproduction number of EVD2 to values less than the ratio 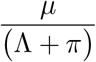 contributes to eradicate the strain from the population and values of *R*_2_ greater than the ratio leads to an infinite number of individuals infected by EVD2. But the time duration of two consecutive strains is not constant in this case, and it is uncertain how to determine the number of impulses that will help to limit the number of individuals infected by EVD2. We then introduce a fixed impulse period *τ* = *t*_*k*+1_ − *t*_*k*_ and obtain from equations (21),

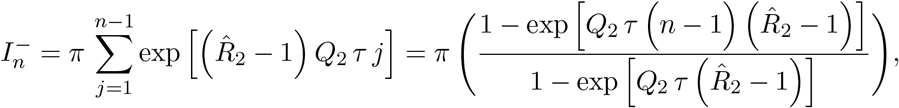

and

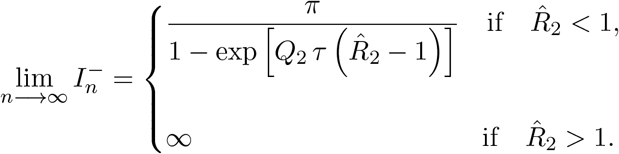

So, the number of individuals infected by EVD2 is bounded if 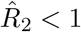 and reducing *R*_2_ to values less than 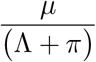 helps to limit the spread of EVD2, but not to clear it from the population. If the total population size is constant (Λ + *π* = *µ*), then 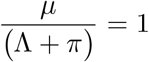 and reducing *R*_2_ to values less than one will slow down the spread of EVD2. The maximum number of individuals infected by EVD2 is then

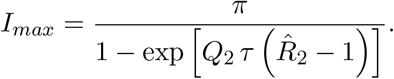

Limiting the number of individuals infected by EVD2 to *I*_*max*_ is equivalent to 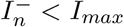 which implies that *τ > τ*_*min*_ with

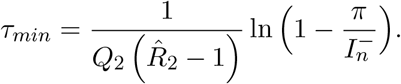

The time lag between two impulses should then be greater than the minimum period *τ*_*min*_ if one wants the maximum number of individuals infected by EVD2 to be *I*_*max*_.

Between two consecutive impulses, the model dynamics is similar to the model with mutation of the resident strain

## 5 Model of Ebola dynamics with mutation of the resident strain

Ebola virus glycoprotein with increased infectivity dominated the 2013 − 2016 epidemic [25]. Viral mutations of Ebola virus occurred over successive human-to-human transmission, improving the adaptation of the virus to the human host and making it more infectious [25]. To understand EVD dynamics with a viral mutation, we consider a scenario where the resident strain (strain 1) mutates and gives rise to strain 2. If we set *σ* = *π* = 0 in the flow diagram in Figure 1, the flow of individuals between the different compartments of the model is represented by the system of equations (22)-(26).

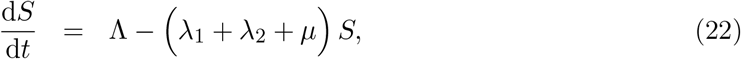

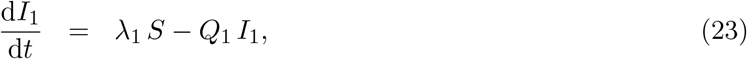

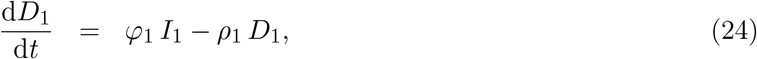

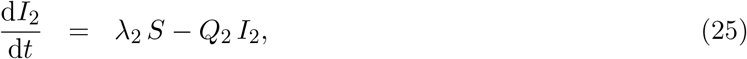

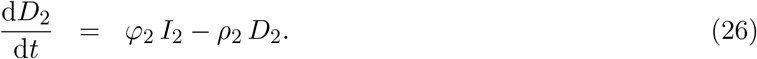

### Theorem 5.1.

The solutions of the system of equations (22)-(26) exist and are non-negative for non-negative initial conditions. The invariant set is

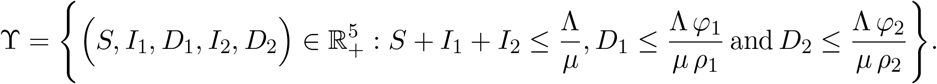

*Proof*. The proof follows that of Theorem 3.2.

### 5.0.1 Strain 2 free equilibrium

Considering a mutant strain, it is important to note that just before the mutation, EVD1 is the only existing strain in the country. In this case, *R*_0_ = *R*_1_ and the endemic equilibrium *Ê*_1_ is given by 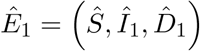 where

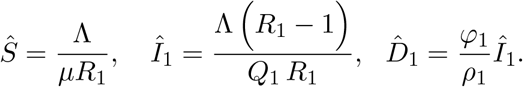

#### Theorem 5.2.

The endemic equilibrium *Ê*_1_ exists for *R*_1_ *>* 1 and is locally asymptotically stable.

*Proof*. As the mutation goes on, EVD2 can invade EVD1 when the latter is at equilibrium and the invasion reproduction number is given by

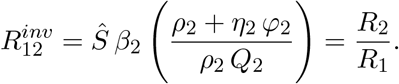

The use of the next generation matrix method to compute 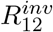 guarantees the local stability of the system at *Ê*_1_.

Figure 3 illustrates the coexistence of the resident and the mutated strain. Figure 3(a) shows the case where the two strains are equally infectious and we observe a rapid decrease of the number of infected humans for both strains from the 8*th* month and EVD free equilibrium is reached after 18 months for the chosen parameter values. This is due to a severe competition between the two strains for the susceptible population. Figure 3(b) illustrates the case where the mutated strain is more infectious than the resident one. The competition for the susceptible population is won by the most infectious strain, which remains endemic until the 20*th* month, while the resident strain dies out after 16 months for the hypothetically chosen parameter values. A viral mutation is often source of complications for disease control as it demands more research to understand the pattern of the new strain. In the case of EVD, if the mutation of a strain does not change its degree of infectivity, then control measures aiming at eradicating the mutated strain can be similar to those used to stop the resident strain as both strains present the same dynamics over time as shown in Figure 3(a). However, control measures must be adapted to the level of infectivity of the mutated strain if its severity is different from the one of the resident strain as it was the case during the last outbreak of 2013 − 2016, during which health authorities had to adapt the control measures to the high infectivity of Zaire ebolavirus strain. More intensive and efficient control measures like a faster contact tracing, a larger educational campaigns, more quarantines and hospitalisations of infected individuals, must be implemented in case of a more severe EVD epidemic.

**Figure 3:**
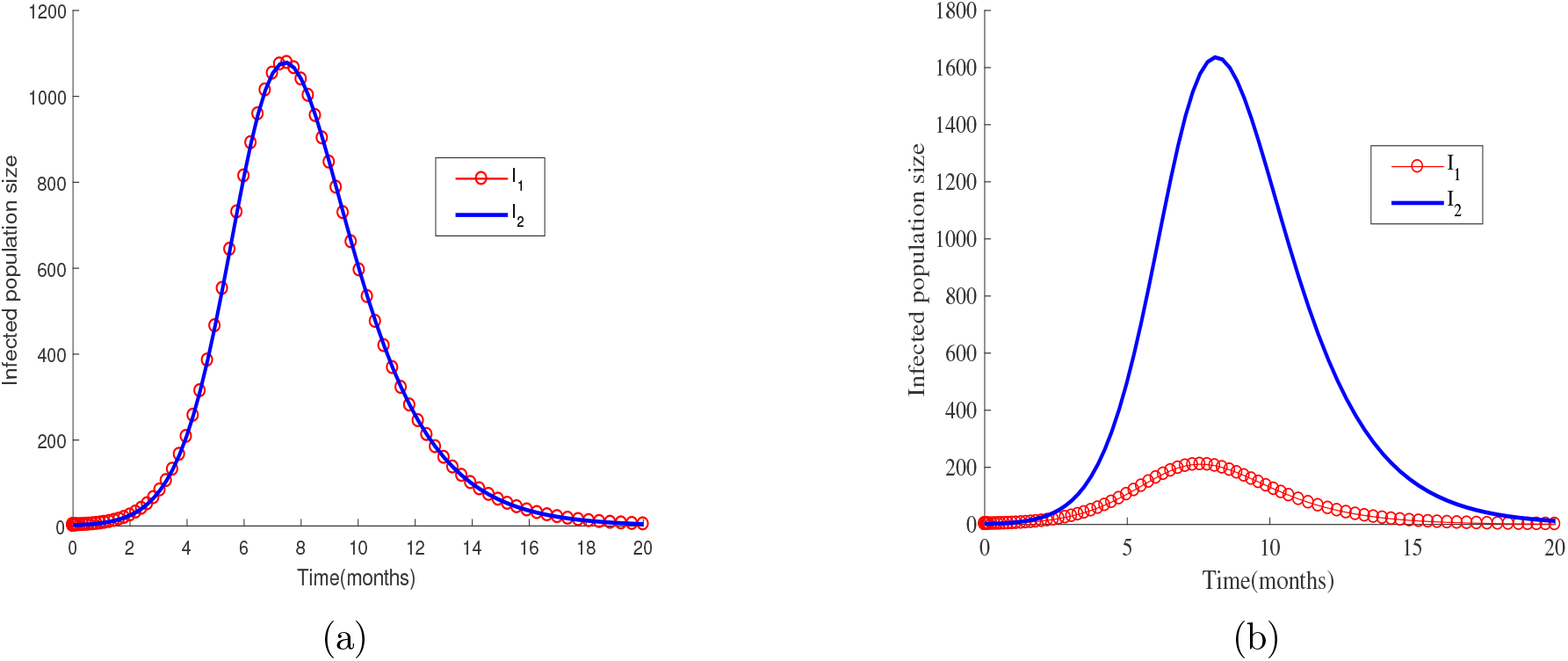
Dynamics of the number of EVD infected individuals when the resident strain mutates. The parameters used are same as those in Figure 4 with *β*_1_ = *β*_2_ = 9 × 10^−5^ in (a) and *β*_1_ = 7 × 10^−5^, *β*_2_ = 9 × 10^−5^ in (b).

### 5.0.2 Coexistence equilibrium

The steady state 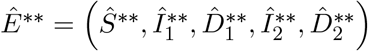 is the solution of system (22)-(26) with

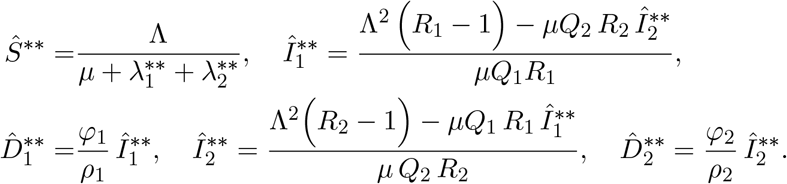

*Ê*\* * is the unique endemic equilibrium point in this case. *R*_1_ *>* 1 is a sufficient condition for strain 1 to exist at an endemic state in the absence of mutation. But in case of mutation, this condition becomes necessary but not sufficient for the two strains to coexist because of the competition for the susceptible population.

## 6 Numerical simulations

Borders in Africa are porous in general, giving rise to the possibility of a continuous migration of populations. Seasons and poor economic conditions motivate frequent movements across borders, which are similar to impulsive movements of populations. For illustrative purposes, we simulate these scenarios in this section, focusing on the effect of increased immigration rate of infectives, increased immigration frequency of infectives and increased infectivity of Ebolavirus strains.

We simulate a scenario where an impulsive immigration of infectives introduces strain 2 is in a country already affected by strain 1. We consider a fixed period of impulse *τ* during which *n* impulses occur. We vary the immigration rate of individuals affected by EVD2 and the period of the impulses is varied as well. The results are given in Figures 4(a) and 4(b). In both figures, we observe that the number of individuals infected by EVD1 is larger when the immigration rate of those infected by EVD2 is lower. This is due to the competition between the two strains for the susceptible population, which is more advantageous for EVD1 in this case. The number of individuals infected by EVD2 on the contrary is larger when *π* is increased. This is an expected result since the number of individuals infected by EVD2 is first fed by the immigration of infectives.

**Figure 4:**
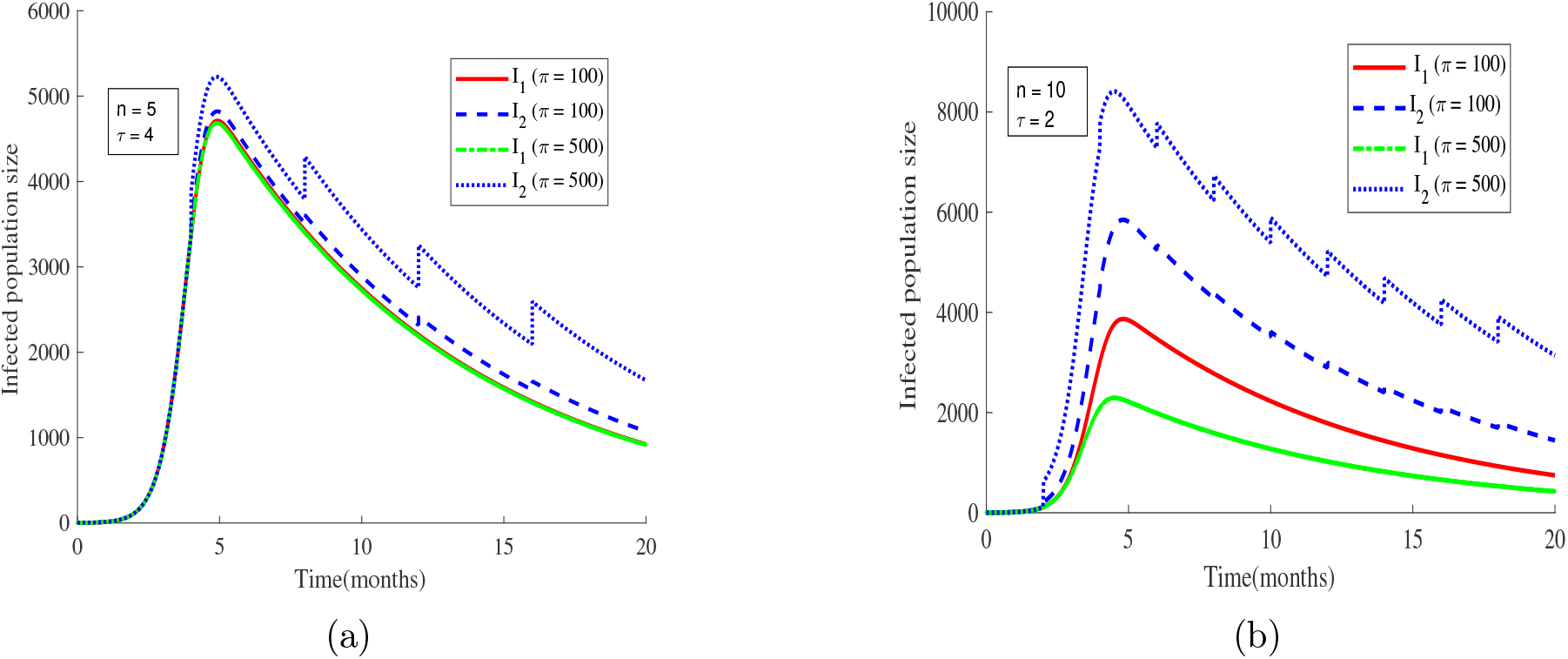
Evolution of the number of EVD infected individuals. The parameters used are Λ = 8.37, *µ* = 0.1, *β*_1_ = *β*_2_ = 9 × 10^−5^, *α*_1_ = *α*_2_ = 0.012, *η*_1_ = *η*_2_ = 2.5, *φ*_1_ = *φ*_2_ = 0.5, *ρ*_1_ = *ρ*_2_ = 0.9

The number of impulses during a fixed period affects the extent of EVD epidemic. In Figure 4(a), we have 5 impulses within 20 months whereas in Figure 4(b) we have 10 impulses within the same period. Irrespective of the strain, the maximum number of infected individuals is 5500 in Figure 4(a) and 8500 in Figure 4(b). Increasing the frequency of the impulses has thus increased the number of infected individuals. The maximum number of individuals infected by EVD2 is reached a bit earlier when the number or frequency of the impulses is increased. Delayed and less intensive immigration of infectives is then advocated. Although a reduced number of impulses allows more infections due to EDV1, a total eradication of EVD2 gives the possibility of reaching a globally stable DFE. The formulation of control measures to eradicate EVD1 is easier in this case. Because road boundaries in Africa are porous, we advocate for more educational campaigns, economic development and good governance so as to limit the movements of populations which is often due to these causes.

We simulate the constant migration of infectives scenario and obtain Figure 5. Figure 5(a) shows that more individuals are infected by EVD2 when the two strains are equally infectious for the chosen parameter values. This result is due to the constant immigration of infectives. In Figure 5(b), we observe that the competition for the susceptibles is favourable to the most infectious strain for about 9 months for the chosen parameter values. From the 10*th* month onwards, we observe that strain 1 reaches the DFE point while strain 2 remains endemic although it is the less infectious strain. This can be explained by the fact that the competition for the susceptible population during the previous months depleted the susceptible population so that EVD1 has a reduced chance of infection. Besides, the recruitment of infectives immigrant is constant and continuously feeds the population of individuals infected by EVD2. This is why EVD2 remains endemic even when there are fewer susceptible individuals to infect. Controls aiming at stopping EVD1 and EVD2 must then also consider the degree of infectivity of each ebolavirus strain. The most infectious strain should be first eliminated as it infects and certainly kills more individuals. Besides, very strict controls at the different entries of a country are necessary to completely stop both strains of Ebolavirus from invading the population.

**Figure 5:**
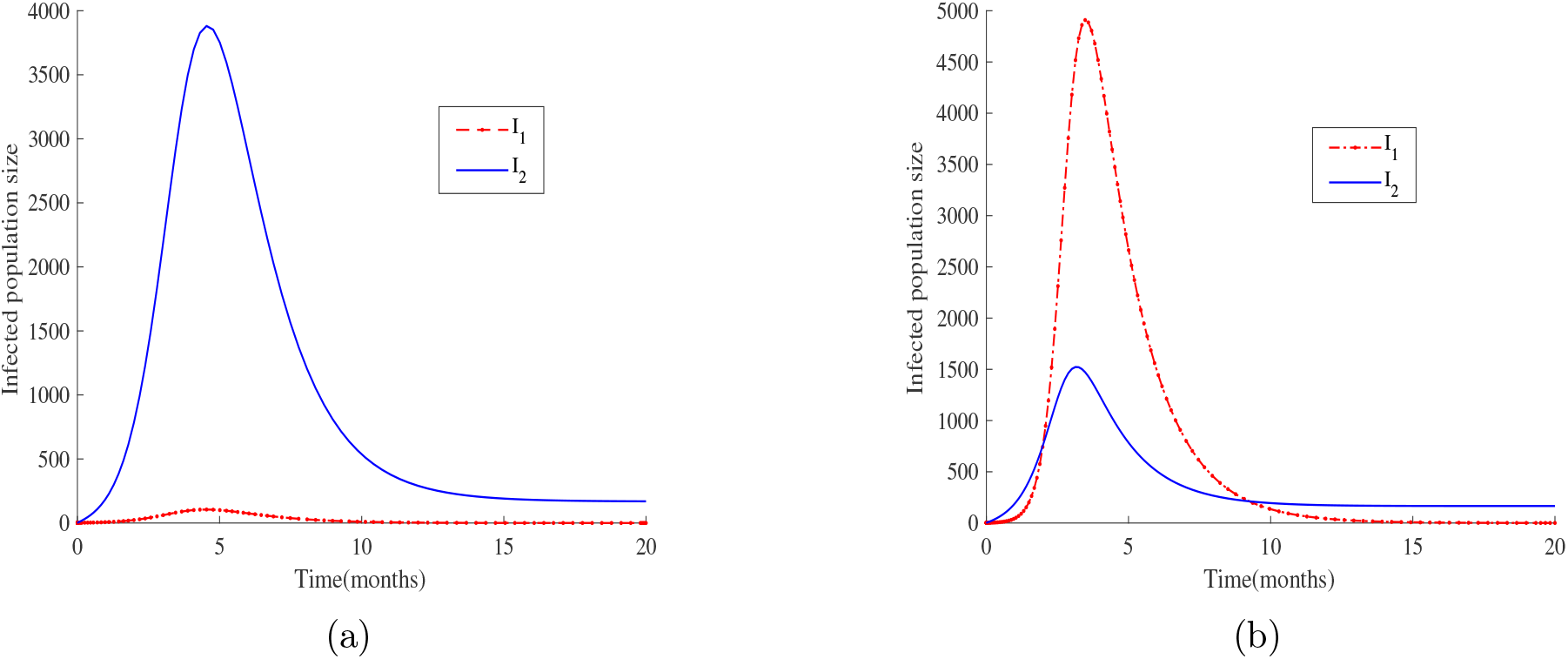
The parameters have the same values as in Figure 4 with *β*_1_ = *β*_2_ = 9 × 10^−5^ in (a), *β*_1_ = 2 × 10^−4^ and *β*_2_ = 9 × 10^−5^ in (b), *π* = 100

The dynamics of EVD represented in Figures 3(a), 4(b) and 5(a) for *π* = 100 shows that, when the same parameter values are used for the simulation, the lowest number of individuals infected by EVD2 (1100) is attained in the case of mutation of the resident strain and the largest number (8900) is reached in the case of impulsive migration of infectives. In Figure 5(a), the highest number of individuals infected by EVD2 is less than 8900. We expected this number to be greater than 8900 since the migration of infectives is without interruption in the case of constant migration. But the large number of infected immigrant transmits EVD2 on a large scale and depletes the susceptible population. This later results in fewer individuals exposed to EVD2. In the case of impulsive migration of infectives, the time lag between two consecutive impulses allows for the susceptible population size to increase and results in higher number of individuals exposed to EVD2 as shown in Figure 4(b). The supplementary source of infectives generated in the case of migration of infectives represented in Figure 4(b) does not exist in the case of mutation of the resident strain and this explains the low number of individuals infected by EVD2 as shown in Figure 3(a). Mutation of the resident strain appears then to be the preferable mean of introduction of a new strain into a population for the chosen parameter values, as it generates less infections. Although the impulsive migration of infectives generates a higher number of infected individuals, it does not deplete rapidly the susceptible population and gives time for control measures implementation. We then suggest that once a new strain is noticed within the boundaries of a country or in his neighbourhood, migration services should privilege impulsive movement to continuous movement of populations at their boundaries. A total closure of the boundaries can be considered if the control cannot be well conducted.

## 7 Conclusion

Movements of infected individuals is a reality and introduced Zaire ebolavirus strain in Liberia and Sierra Leone in 2014 [26]. We formulated and analysed in this paper, a model of EVD dynamics in which EVD2 is introduced in a country where EVD1 is endemic.

First, we considered that the immigration of individuals infected by EVD2 is continuous. The mathematical analysis of the model indicated the existence of a locally stable disease free equilibrium and an endemic equilibrium. It showed that when the invasion reproduction number of EVD2 is greater than one, EVD2 invades the country. The two strains coexist at an endemic state when the reproduction number of EVD1 is greater than the one of EVD2. Numerical simulations indicated a rapid and abrupt increase of the number of individuals infected by EVD2 which allows less time for control measures’ implementation. A fast decrease of the number of individuals infected by EVD1 was noticed as well and this can be the ideal solution if clearing EDV1 from the population is the objective.

Second, immigration of individuals infected by EVD2 was considered to be impulsive and we proved that the reproduction number of EVD2 must be less than the ratio (*µ/* Λ + *π*) in order to limit the number of individuals infected by EVD2. We also found that a fixed period of impulse is better for EVD2 control since it helps to evaluate with precision the number of impulses that minimizes the number of individuals infected by EVD2. Numerical simulations’ results indicated that the smaller the immigration rate of individuals infected by EVD2, the larger the number of individuals infected by EVD1. This sheds the light on the competition between the two ebolavirus strains for the susceptible population.

Finally, we have considered the case where the resident strain of EVD in a country mutates and gives rise to a new one. The number of individuals infected by the new strain is reduced because of the absence of immigration of infectives. The mathematical analysis of the model in this case indicated a competition between the resident and the mutated strains. We found that this competition is favourable to the most infectious strain, just as in the case of a continuous immigration of infectives. Results from the numerical simulations indicated that control measures should be adapted to tackle even the most severe strain.

In summary, we state that the impulsive type of migration of individuals infected by the less infectious EVD strain would be a better scenario. We argue that this type of migration of infectives is manageable because it allows more time for control measures to be implemented, increasing the chances of stopping EVD irrespective of the strain. In case of mutation of an EVD strain, control measures must be adapted to the level of infectivity of the new strain. An unknown outbreak started in Guinea in December 2013 and was only declared as an EVD outbreak later in March 2014 by the WHO [26]. In case of co-infection by different strains of EVD, such delay will lead to a death toll far larger than the 11000 recorded in 2016. An impulsive movement of infectives even in such situation, is preferable to a continuous movement of infectives as it delays the increase in the number of infected migrants. The study presented in this paper considers two of the five existing strains of Ebola virus and describes well the scenario of a multi-strain epidemic of Ebola. Considering more strains is what we endeavour to do in the future as it will give a bigger picture of a potential co-infection situation.

## Data Availability

All data produced in the present work are contained in the manuscript

## Author Contributions

The authors of this manuscript equally contributed to the conceptualization, analysis, simulations and the writing-up of the manuscript.

## Acknowledgements

The authors authors acknowledge the support of their respective institutions in the production of the manuscript.

